# Impact of the COVID-19 pandemic on breast cancer screening indicators in a Spanish population-based program: a cohort study

**DOI:** 10.1101/2022.03.04.22271911

**Authors:** Guillermo Bosch, Margarita Posso, Javier Louro, Marta Roman, Miquel Porta, Xavier Castells, Francesc Macià

## Abstract

**Background:** To assess the effect of the COVID-19 pandemic on performance indicators in the population-based breast cancer screening program of Parc de Salut Mar (PSMAR), Barcelona, Spain.

**Methods:** We conducted a before-and-after, quasi-experimental study to evaluate participation, recall, false-positives, cancer detection rate, and cancer characteristics in our screening population from March 2020 to March 2021 compared with the four previous rounds (2012-2019). Using independent logistic regression models, we estimated the adjusted odds ratios (aOR) of each of the performance indicators for the COVID-19 period, controlling by type of screening (prevalent or incident), socioeconomic index, family history of breast cancer, and menopausal status. We analyzed 144,779 observations from 47,571 women.

**Results:** During the COVID-19 period, the odds of participation were 11% lower in first-time invitees (aOR=0.89[95%CI=0.84-0.96]) and in those who had previously participated regularly and irregularly (aOR=0.65 [95%CI=0.61-0.69] and aOR=0.93 [95%CI=0.85-1.03], respectively). Participation showed a modest increase in women not attending any of the previous rounds (aOR=1.07 [95%CI=0.99-1.17]). The recall rate slightly decreased in both prevalent and incident screening (aOR=0.89 [95%CI=0.78-1.01] and aOR=0.89 [95%CI=0.79-1.00], respectively). No significant differences were observed in false-positives (prevalent - aOR=1.07 [95%CI=0.92-1.24] and incident screening -aOR=0.94 [95%CI=0.82-1.08]), cancer detection rate (aOR=0.91 [95%CI=0.69-1.18]), or cancer stages.

**Conclusions:** The COVID-19 pandemic negatively affected screening attendance, especially in previous participants and newcomers. We found no marked differences in recall, false-positives, or cancer detection, indicating the program’s resilience. There is a need for further evaluations of interval cancers and potential diagnostic delays.

## Introduction

In numerous health systems cancer screening programs were among the first activities interrupted by the COVID-19 pandemic after its irruption in early 2020. As reported in a survey by the International Cancer Screening Network, 97% of participating settings reported that COVID-19 had adversely impacted their screening programs, while 90% partially suspended their activity (1,2). Even in countries with notable success in containing the pandemic, like Taiwan, the population attending screening decreased during the first half of 2020 (3).

In Europe, breast cancer screening is mostly provided through organized programs offering routine mammography examination to women aged from 45-50 to 69-74 years. The programs follow the guidelines of the European Commission Initiative for Breast Cancer Screening and Diagnosis (4). These guidelines recommend an evidence-based set of performance indicators to evaluate the quality of the screening provision (5). The suspension of these programs led to a reduction in cancer diagnoses. For instance, in the Netherlands and Austria, the number of breast cancer diagnoses decreased substantially and remained lower than expected until screening was rebooted (6,7), while in Italy, between January and May 2020, 53% fewer screens were performed, with a median delay of 2.7-month for screening mammograms (8).

Currently, the evidence of the effect of the COVID-19 pandemic on breast cancer screening performance indicators has been mostly provided by simulation models and longitudinal studies are scarce. In Canada, Yong et al. used a mathematical model to estimate that a three-month halt would have led to 664,000 fewer screening mammograms than expected, based on nationwide data from the previous year. It would also have decreased breast cancer diagnoses by 7% in 2020 and caused 110 excess deaths by 2029 (9). Similar models in Italy reported that 8,125 breast cancer diagnoses were expected to be delayed due to a three-month interruption of screening programs, representing 25% of the 32,500 yearly screening diagnoses nationwide (10).

Spain was one of the first and most affected countries in Europe during the spring of 2020 (11,12). On March 14th, a general lockdown was enforced, and breast cancer screening was interrupted (13). Restrictive measures were slowly withdrawn during the following three months until June 21^st^, when the lockdown ended (14). To reintroduce the screening programs as soon as possible while continuing to control the risk of COVID-19 transmission, mammography centers established new safety guidelines (15,16).

Given the scarcity of longitudinal studies, we used a before-and-after design including data from a population-based program from 2012 to 2021. We aimed to assess the impact of the COVID-19 pandemic on the performance indicators of the program of Parc de Salut Mar (PSMAR) of Barcelona, Catalonia, Spain.

## Methods

### Study design

In this before-and-after study, we compared the population-based breast cancer screening indicators obtained in a single population before and after the COVID-19 pandemic.

### Study population

In Spain, publicly funded mammographic screening for breast cancer is offered every two years to women aged 50 to 69 years (17). The screening examination at PSMAR consists of both a mediolateral oblique and a craniocaudal digital (two-dimensional) mammographic view of each breast. Two independent radiologists with extensive experience perform blinded double reading of mammograms. Disagreements are resolved by a third senior radiologist (18). The program covers the population of four districts of the city of Barcelona, with around 620,000 inhabitants. Until the current pandemic, screening invitations were sent by postal mail with a pre-scheduled mammography appointment. Since June 2020, previous participants have also received a reminder by telephone.

Invitations are issued during the two-year duration of each screening round according to the geographical criteria set by Basic Health Areas (BHA), which are the basic territorial healthcare units of the city. In this analysis, we used data from 10 out of the 25 BHA covered by the PSMAR breast cancer screening program. The 10 BHA selected were those affected by the interruption and delay of the screening program during the first year of the pandemic, invitations for women from such BHA should have been sent out between March 2020 and March 2021.

For this study, the pre-COVID-19 period started in March 2012, ended in March 2019, and was divided into four pre-COVID-19 rounds of two years each. The post-COVID-19 period, therefore, went from March 2020 to March 2021, and included one screening round. We extended the follow-up until September 2021 to include the process of cancer diagnosis for women attending screening in the post-COVID round.

We obtained a total of 144,779 observations, from 47,571 eligible women throughout the 10 years of study. Each of these observations represented an invitation to the screening program. In our study population, age group, socioeconomic status, and type of screening round were statistically different in the post- and pre-COVID-19 periods, (Table 1). The percentage of invited women living in high-income areas decreased slightly (−1.03%) as did that of women younger than 55 years (−1.83%). The distribution of the type of screening of invited women also changed, with a higher percentage of invitations for prevalent screening (+1.69%), especially first-time invitees (+2.90%).

**Table 1.**
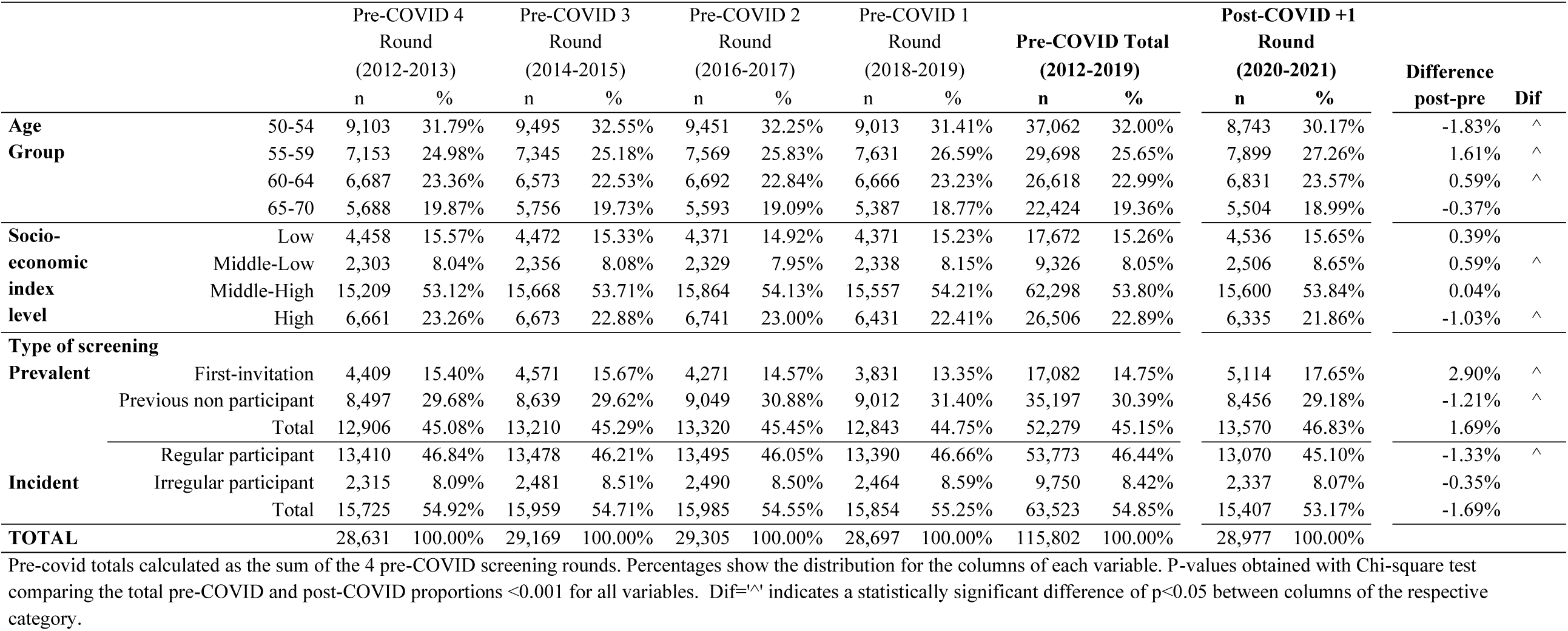
Baseline characteristics of women invited to the PSMAR breast cancer screening program in the BHA (Basic Health Areas) affected by the COVID-19 pandemic in the area of reference of Parc de Salut Mar (PSMAR), in Barcelona, Spain in the period 2012-2021 by screening round.

### Data sources

All data from women eligible for screening was obtained from the management database of the program, which is updated yearly with the screening status and baseline characteristics of both participants and non-participants over the years. We completed the information on cancer histology and tumor stages with data the clinical and pathological records.

Each screening invitation was considered an independent measurement. All measurements were pseudo-anonymized by using an individual ID for each woman while removing all personal data. Therefore, although multiple measurements per woman could be obtained from invitations in different rounds, all of them shared the same pseudo-anonymized ID.

### Outcomes

From the screening database of the program, we obtained the indicators on the selected BHA for the 2020-21 screening round, which, as mentioned, was categorized as the post-COVID-19 period. We compared the post-COVID-19 indicators with those from the four previous screening rounds of the same BHA, categorized as the pre-COVID-19 period. We used four main indicators of the program: participation, recall, false-positives, and detection rate. In addition, we also compared the following characteristics of the detected tumors, histology (invasive vs. in situ), tumor size, lymphatic invasion, the presence of metastases, and stage at diagnosis.

Participation was measured as the percentage of women invited for screening who underwent mammography in the corresponding round. Invited women were those fulfilling the selection criteria (age 50 to 69 years, residence in the selected BHA) and who did not meet of the exclusion criteria of the program. The main exclusion criteria were a change of address outside the geographic area of the program, previous breast cancer, high hereditary risk of breast cancer and errors in identification or personal data.

The other three outcomes were analyzed using only the screening participants. The recall rate was estimated as the percentage of participants who were advised to undergo further assessment to rule out malignancy, whether non-invasive or invasive (ultrasound, tomosynthesis, contrast-enhanced mammography, biopsy, and/or others). False positives were estimated based on the percentage of women who underwent additional non-invasive or invasive assessments but who did not have a diagnosis of cancer after completion of additional examinations. The detection rate was the number of breast cancers detected at screening per 1000 participants. We calculated this rate, stratifying by type of breast cancer histology (i.e., the invasive or in situ cancer detection rate).

We stratified all the invitations by type of screening between prevalent or incident screening. Prevalent screening refers to the process of inviting women who have never participated in screening, while incident screening refers to inviting previous participants. In terms of prevalent screening, we differentiated between first-time invitees, and non-participants, referring to previously invited women who had never participated. For incident screening, we differentiated between previous participants who had participated in the previous round (regular participants) and those not participating in the last round (irregular participants).

We categorized women according to their age at the time of invitation in four groups: 50-54, 55-59, 60-64, and 65-70 years old. Socio-economic status was estimated with a compound socioeconomic index, created by the Government of Catalonia to assign resources to primary health care, based on the index of each BHA (19). Each woman was assigned the socioeconomic index of the BHA where she was living. Higher values denote a lower socioeconomic level.

We evaluated clinical variables such as menopausal status and family history of breast cancer. Breast cancer histology differentiates between in situ and invasive tumors. According to the TNM Breast Cancer 8^th^ Edition classification (20), tumor size was measured in millimeters, lymphatic invasion as the extension of malignant cells, metastasis as its presence or absence, and stage at diagnosis as I, II, III, and IV TNM categories. We used the pathology (p)TNM preferably, and only used the clinical (c)TNM for women with neoadjuvant treatment (21). Other epidemiological and clinical variables such as educational level or history of hormone replacement therapy were not included in the analyses due to a high percentage of missing values (>10%).

### Statistical analysis

We first compared the characteristics of the invited population among the different screening rounds to describe variations in their distribution. We evaluated differences in the categories using the chi-square test or the exact Fisher test when appropriate.

Then, we created independent logistic regression models to estimate adjusted odds ratios (aOR) of each of the performance indicators and their corresponding 95% confidence intervals (95% CI) for the COVID-19 period, adjusting by the clinically relevant variables.

For participation, we included the following variables in the model: type of screening round (prevalent vs. incident), age group, and socio-economic index. We found a strong interaction between COVID-19 and the type of screening round. Therefore, we created a new variable, which represented this interaction. Hence, the final models for participation differentiated four screening groups (prevalent-first-time invitee, prevalent-previous nonparticipant, incident-regular participant, and incident-irregular participant). We obtained crude results and adjusted by age and socioeconomic index.

We created three additional models, including only participants, to assess the impact of COVID-19 on the other main indicators of the screening program: recall, false-positives, and the screen-detected cancer rate (invasive or in situ). These models were adjusted for age group, menopausal status, and breast cancer family history.

Finally, we compared the stage at diagnosis and the remaining cancer characteristics (size, lymph node invasion, and metastasis invasion) of cases detected in the screening program in the pre- and post-COVID-19 periods.

Statistical tests were two-sided and all p-values lower than .05 were considered statistically significant.

SPSS version 25 software was used for the creation and validation of the database and recodification of variables, while statistical software R version 3.5.0 (Development Core Team, 2014) was used for the logistic regression models.

### Ethical aspects

The study guaranteed Spain’s legal regulations on data confidentiality (law 15/99 of December 13 on the protection of personal data). Due to the retrospective nature of the study and the absence of direct contact with women, which did not affect their relationship with the program, informed consent was waived by the Ethics Committee of PSMAR, which approved the study (reg.2021/9866).

## Results

The participation in the program was affected differently depending on the type of screening. The odds of participating during the post-COVID-19 period were 10% lower (aOR=0.90 95% CI-, 0.84-0.96) for the group of first-time invitees than during the pre-COVID-19 period. The aOR was 1.07 (95% CI, 0.99-1.17) for the previous non-participant group between the post- and pre-COVID-19 periods. For the group of women who had participated in the previous round (regular participants), the aOR of participation was 0.65 (95% CI 0.61-0.69), and for those not participating in the last round (irregular participants), the aOR was 0.93 (95% CI 0.85-1.03) (Figure 1).

**Figure 1.**
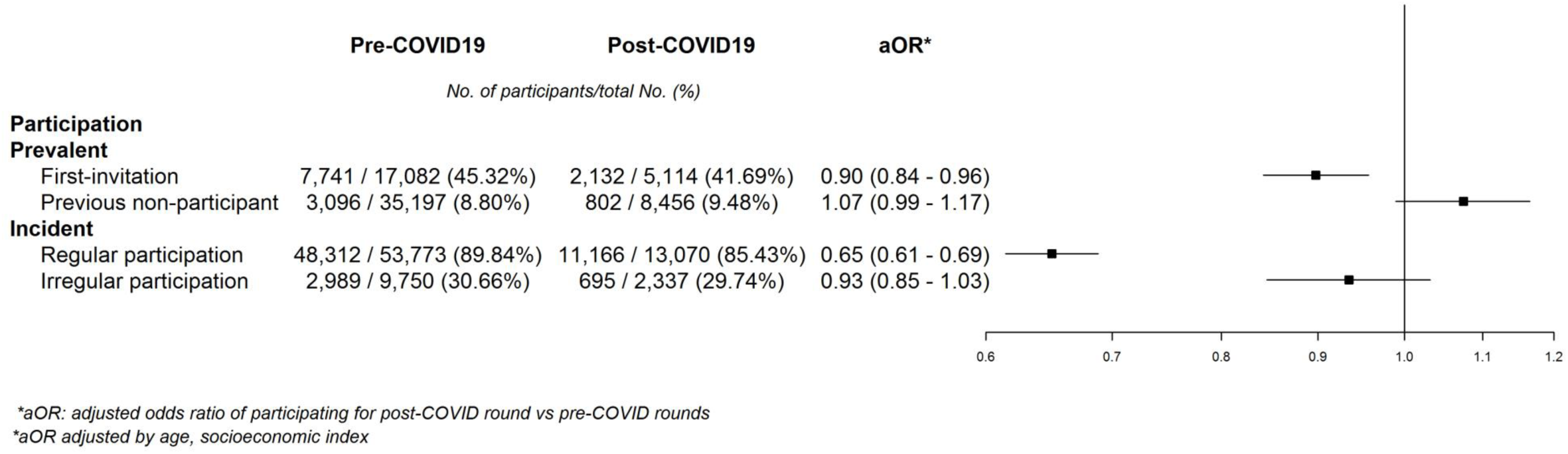
Adjusted Odds Ratios of Pre-post COVID-19 models for participation.

We also found statistically significant differences in the distribution of baseline characteristics of participants during the post-COVID-19 round compared with the mean distributions of the pre-COVID-19 period (Table 2). The percentage of participants younger than 55 years decreased (−1.64%) but the percentage aged between 55 and 59 years increased (+1.73%). The percentage of participants from high socioeconomic level areas slightly decreased (−0.62%). The biggest changes in the distribution of participants were seen between types of screening, with a substantial decrease among participants in the incident screening group (−2.39%) and an increase in the percentage of prevalent screening, especially first-time invitees (+1.96%). The percentage of participants with a family history of breast cancer increased by 2.53%.

**Table 2.**
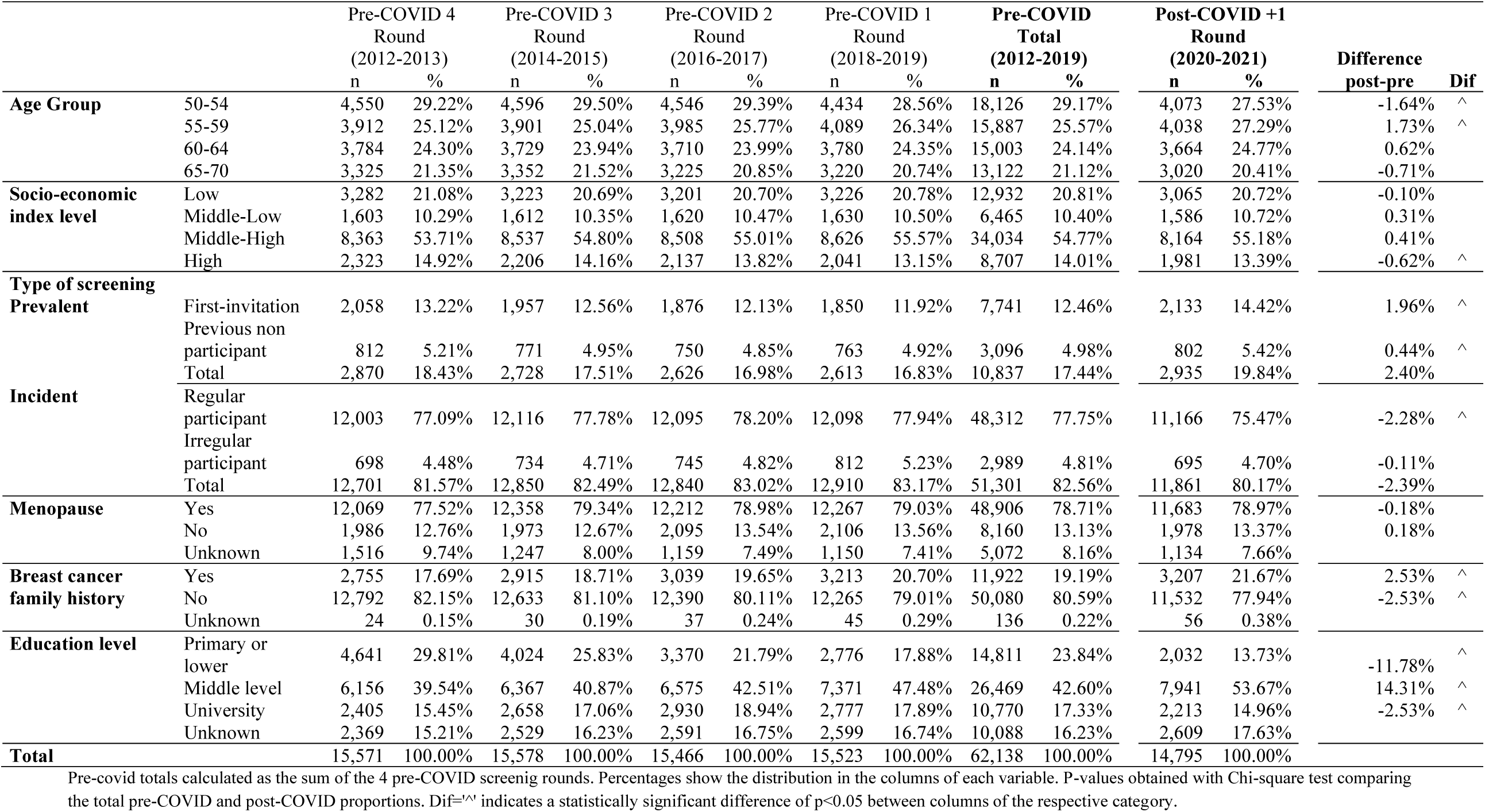
Baseline characteristics of participants of the PSMAR breast cancer screening program in the BHA affected by the COVID-19 pandemic in the area of reference of Parc de Salut Mar (PSMAR), in Barcelona, Spain in the period 2012-2021 by screening round.

The analysis of recall revealed modest decreases of 11% in the likelihood of being advised to undergo additional testing the post-COVID-19 period in both the prevalent and the incident screening groups (aOR 0.89 [95% CI 0.78-1.01], and aOR 0.89 [95% CI 0.79-1.00]). The aOR of a false positive result for prevalent and incident screening was 1.07 (95% CI, 0.92-1.24) and 0.94 (95% CI, 0.82-1.08), respectively. The aOR of cancer detection in the post-COVID vs the pre-COVID-19 period was 1.01 (95% CI 0.56-1.71) and 0.87 (95% CI 0.63-1.17) in the prevalent and incident screening groups, respectively (Figure 2).

**Figure 2.**
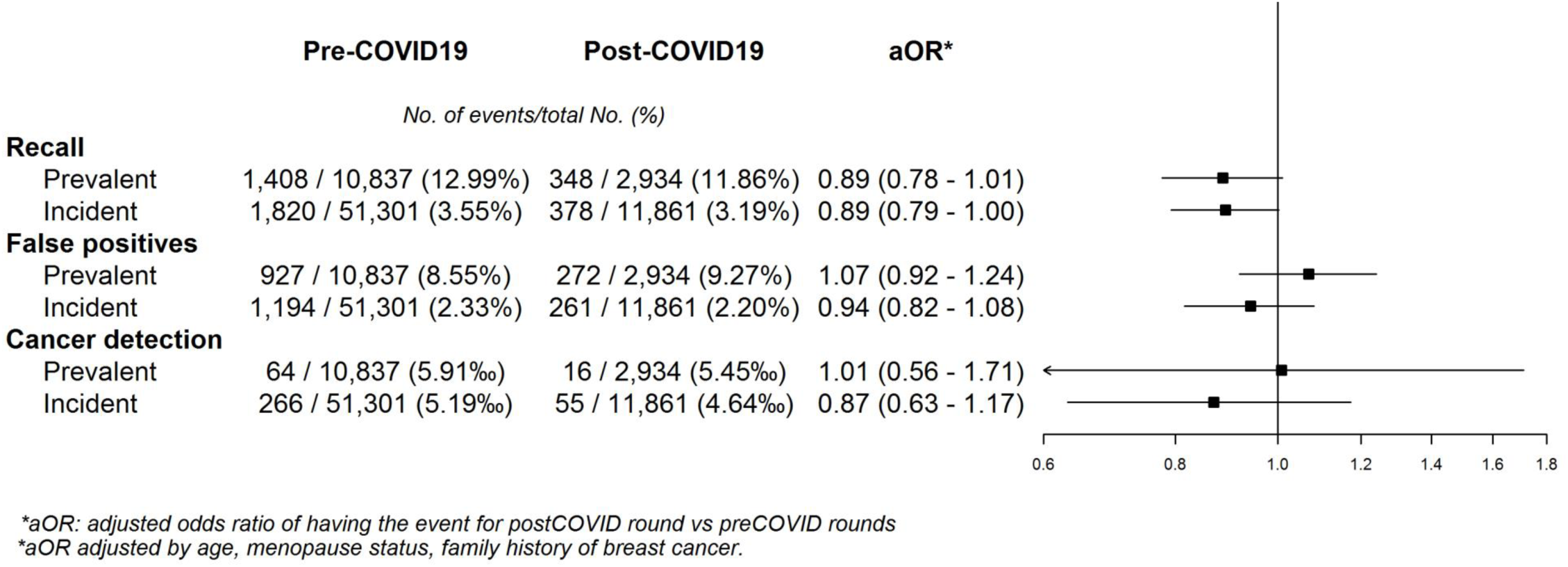
Adjusted Odds Ratios of Pre-post COVID-19 models for recall, false positives, and cancer detection in participants.

When differentiating between cancer histology, we found no statistically significant reductions in the odds of being diagnosed with a carcinoma in situ (aOR 0.74 [95% CI 0.32-1.47]) or an invasive tumor (aOR 0.95 [95% CI 0.70-1.26]), whereas the aOR for all tumors was 0.91 [95% CI 0.69-1.18] (Figure 3). Finally, we observed no statistically significant differences in the distribution of the tumor size, lymphatic invasion, the presence of metastasis or stage at diagnosis between the pre- and post-COVID-19 periods. A statistically non-significant decrease of 4.47% in in-situ tumors, and a non-significant increase of 4.95% in stages I were noted (Table 3).

**Figure 3.**
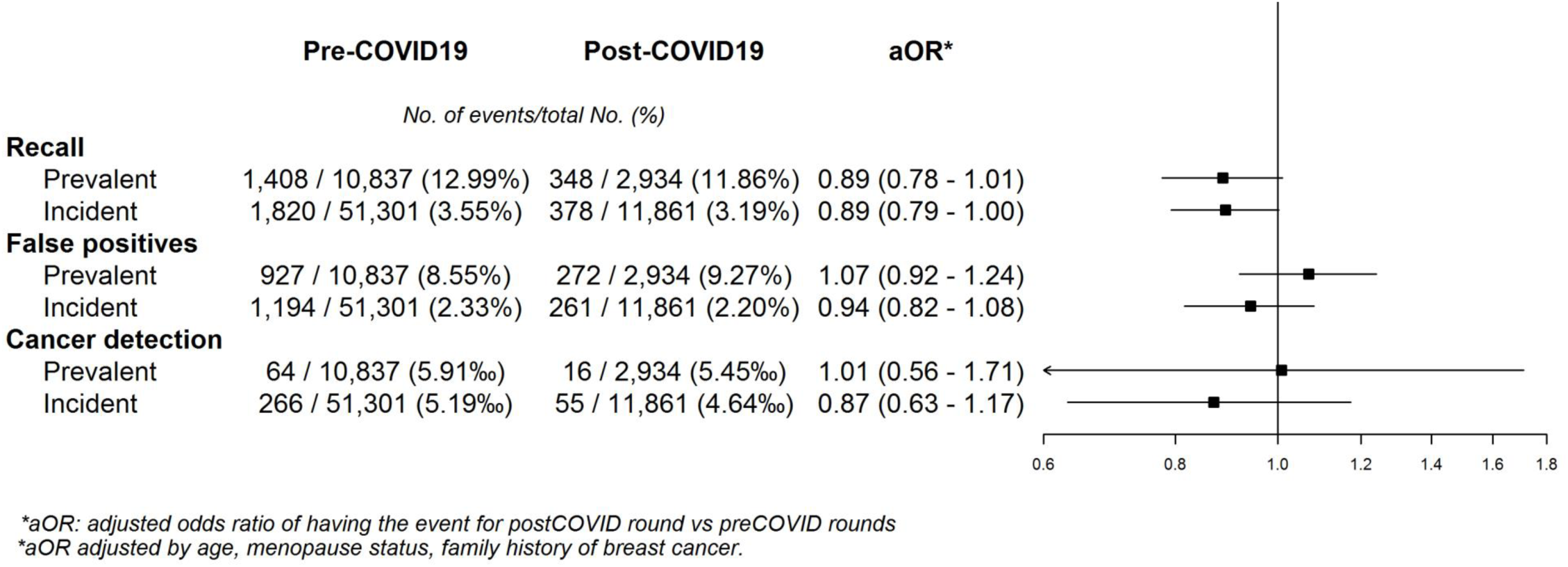
Adjusted Odds Ratios of Pre-post COVID-19 models for cancer detection according to histology.

**Table 3.** Staging of cancers detected in each screening round in the BHA affected by the COVID-19 pandemic in the area of reference of Parc de Salut Mar (PSMAR), in Barcelona, Spain in the period 2012-2021 by screening round.

|  |  | Pre-COVID Total<br>(2012-2019) |  | Post-COVID +1 Round<br>(2020-2021) |  | Difference post-pre |
| --- | --- | --- | --- | --- | --- | --- |
|  |  | n | % | n | % |  |
| **T** | In Situ | 51 | 15.45% | 8 | 11.27% | -4.76% |
|  | 1 | 210 | 63.64% | 47 | 66.20% | 0.48% |
|  | 2 | 43 | 13.03% | 13 | 18.31% | 4.92% |
|  | 3 | 8 | 2.42% | 2 | 2.82% | 0.32% |
|  | 4 | 3 | 0.91% | 0 | 0.00% | -0.95% |
|  | Unknown | 15 | 4.55% | 1 | 1.41% |  |
| **N** | 0 | 254 | 76.97% | 58 | 81.69% | 1.97% |
|  | 1 | 47 | 14.24% | 10 | 14.08% | -0.68% |
|  | 2 | 8 | 2.42% | 2 | 2.82% | 0.31% |
|  | 3 | 5 | 1.52% | 0 | 0.00% | -1.59% |
|  | Unknown | 16 | 4.85% | 1 | 1.41% |  |
| **M** | 0 | 307 | 93.03% | 68 | 95.77% | 0.97% |
|  | 1 | 3 | 0.91% | 0 | 0.00% | -0.97% |
|  | Unknown | 20 | 6.06% | 3 | 4.23% |  |
| **Clinical Stage** | In situ | 50 | 15.15% | 8 | 11.27% | -4.47% |
|  | I | 175 | 53.03% | 42 | 59.15% | 4.95% |
|  | II | 62 | 18.79% | 16 | 22.54% | 3.40% |
|  | III | 18 | 5.45% | 2 | 2.82% | -2.90% |
|  | IV | 3 | 0.91% | 0 | 0.00% | -0.97% |
|  | Unknown | 22 | 6.67% | 3 | 4.23% |  |
| **TOTAL** |  | 330 | 100.00% | 71 | 100.00% |  |

Pre-COVID totals calculated as the sum of the four pre-COVID rounds. Difference post-pre calculated comparing the post and the total pre-COVID percentages. Percentages show the distribution in the columns of each variable. P-value of the distribution of each variable calculated with the exact test of Fisher. Dif='^' indicates a significant difference of $p > 0.05$ between columns of the respective category

All crude results did not significantly differ from those adjusted (supplementary file 1).

## Discussion

In this longitudinal study, we found that the pandemic reduced participation, but that this impact differed according to each woman’s history of participation. Although attendance was lower during the pandemic period, we found no significant differences in other performance indicators, such as the frequency of recall for additional tests after mammography, the percentage of false-positive results, or the cancer detection rate.

Women who became eligible for invitation to our population-based screening program for the first time in the post-COVID-19 period were significantly less likely to participate during the pandemic. This effect was also noted, and in a much higher degree, in women who had participated in the previous round. A reduction in participation, although non-statistically significant, was also seen in women with previous irregular participation. Even though we could not identify the exact reasons behind the lower participation, we hypothesize that possible factors could be general insecurity related to attending hospitals, governmental restrictions of movement, fear of COVID-19 infection, and other uncertainties about the safety of participating in the screening process. This hypothesis is supported by data from a Danish study reporting that two of the reasons for postponing or canceling mammography appointments during the first year of the pandemic were fear and lack of clear guidance on the safety of screening (22).

Women who had been previously invited but had never attended our screening invitation seemed to participate slightly more during the pandemic period, although this difference was not statistically significant. The increase in participation was not expected since this group of women is that with the lowest participation in our setting (23). However, this change could be explained by a plausible modification in attitudes to screening with a possible increase in health-consciousness promoted by the pandemic, prompting women who had never been interested in screening to participate for the first time.

Overall, our findings on participation adjusted by age and socioeconomic status showed that the effect of the pandemic on screening attendance depended on each woman’s previous participation status. Although the aim of our study was not to evaluate the factors associated with participation, we found a lower representation of high-income women in the post-COVID-19 period. This observation could be related to the pandemic situation, and should be confirmed, and is contrary to previous evidence. Nevertheless, it may well reflect a real phenomenon. A systematic review of studies conducted before the pandemic reported lower participation in low-income groups, immigrants, non-homeowners, and women with a previous false-positive result (24). Furthermore, studies recently published in the US have reported a decrease in participation, especially in underserved ethnic groups, with lower socioeconomic status, lack of insurance and longer travel time (25,26). Monitoring this information would allow programs to make efforts to promote participation among women at higher risk of not participating, especially under disruptive situations.

Despite the lower participation, the remaining performance indicators in our program did not seem to be significantly affected by the pandemic. Our results showed a non-statistically significant reduction in the recall rates of both prevalent and incident screening. These findings could be due to the increased workload caused by COVID-19 patients at our and many other hospitals, which strongly affected the radiology department in 2020 (27). Regarding the frequency of false positives, we found no significant variation due to the pandemic, suggesting that the diagnostic accuracy of the radiologists reading the mammograms was not materially affected. Similar pieces of evidence of the resilience of our public health-care system have been recently reported in other hospitals in Barcelona (28), suggesting the strong resilience of health professionals working in critical situations. The COVID-19 pandemic has proved to be a stress test for health care systems around the world and the main elements related to highly effective responses have been associated with adaptation of health systems’ capacity, reduction of vulnerability, preservation of health care functions and resources, and activation of comprehensive responses (29).

We found no differences in the odds of screen-detected cancer for either prevalent or incident screenings when comparing the pre- and post-COVID-19 periods. In contrast to our statistical approach to estimate the cancer detection as the number of tumors per 1,000 participants, when the absolute number of diagnoses during the interruption of the screening programs was compared with previous periods, an evident reduction was observed. A study performed in Málaga (Spain) reported that the breast was one of the cancer sites showing a larger decline in cases in April 2020 compared with April 2019. The authors of that study stated that this decline could be explained by the interruption of the screening program (30). Similar results have been found in studies from the Netherlands, Austria, and the United Kingdom (6,7,31).

It is still unknown whether the target strategies to reduce the back-log of women who missed screening due to the pandemic, such as contacting them by telephone calls to schedule an appointment, will help to detect cancers missed during screening disruption. The possible influence of the delay on stage at diagnosis needs further evaluation. Although we found no statistically significant differences between pre- and post-COVID-19 periods in our small sample, we did find a small 5% increase in cases diagnosed at stage II. Similarly, an increased risk of late-stage breast cancer was observed in a month-by-month comparison in Israel in the period following the interruption and restoration of the screening activity (32). Further investigation on stage at diagnosis is essential, especially considering the potential increase in interval cancers due to the delay in the planned mammogram schedule. Moreover, the reduction in participation could increase cancer detection and stage at diagnosis in the next screening round among women who skipped the post-pandemic round, leaving a span of four years between consecutive screenings.

Our study has some limitations. First, we used pseudo-anonymized data and analyzed each invitation as an independent measurement, although a single woman can have more than one invitation. Nevertheless, we considered four groups of type of screening rounds to evaluate attendance, mitigating the fact that the measurements were not truly independent, although this study treated them as though they were. Second, the number of cancers detected during the pandemic period was relatively low, which limited the statistical power of our results.

Our study also has some strengths. To our best knowledge, most of the observational evidence assessing the effect of the pandemic compared screening indicators with the previous year (33–35). In contrast, we included a long period of four previous rounds (eight years) of invitations for the same target population. We took this longitudinal approach since it is known that there are fluctuations in participation and cancer detection that may depend on time (36). Therefore, our approach provides information on the pandemic beyond these common fluctuations

In conclusion, our findings suggest that the impact of the pandemic on screening attendance depends on the type of screening, with women who regularly participate being the most affected. Targeting this specific population with a proactive invitation could be a way to ensure the historically higher participation in this group. Despite this, we should not forget other groups that attended screening less frequently. Our program has proved to be resilient, reducing recall and maintaining invitations, false positives, and the cancer detection rate stable. These results suggest that the roll-out of the program was successful under the stressful situation provoked by the pandemic. Further prospective research is necessary to assess whether other factors played a role in participation during the pandemic, as well as to better characterize the impact of delays on stage at diagnosis and the incidence of interval cancers.

## Data Availability

Source data form all tables and figures can be found in the following Dataset: BOSCH, GUILLERMO, 2022, "Breast cancer screening program invitations (2012-2021)", https://doi.org/10.7910/DVN/VVQNWM, Harvard Dataverse, V1, UNF:6:CaW3sEp4tMsg13z2I1eZbQ== [fileUNF] Data from "Impact on covid19 dataset invited women.sav" was used in tables 1 and 2 and figures 1,2 and 3. Data from "cancer characteristics database.tab" was used in table 3.

## Abbreviations

aOR: adjusted odds ratio
BHA: basic Health Area
CI: Confidence interval
PSMAR: Parc de Salut Mar

## Availability of data and materials

Source data from all tables and figures can be found in the following Dataset: BOSCH, GUILLERMO, 2022, “Breast cancer screening program invitations (2012-2021)”, https://doi.org/10.7910/DVN/VVQNWM, Harvard Dataverse, V1, UNF:6:CaW3sEp4tMsg13z2I1eZbQ== [fileUNF] Data from “Impact on covid19 dataset invited women.sav” was used in tables 1 and 2 and figures 1,2 and 3. Data from “cancer characteristics database.tab” was used in table 3..

## Authors’ contributions

All authors conceptualized and designed the study. Data was collected by GB, MP, and FM. GB, JL, and MR performed the statistical analyses. All authors collaborated in the interpretation of the results. GB, and MP drafted the manuscript. GB, MP, and FM wrote the final version of the manuscript and revised it critically for important intellectual content. All authors read and approved the final manuscript.

## Acknowledgments

we would like to thank Cristina Barrufet, Mercè Esturi and Cristina Hernández in particular for their work and assistance in the performance of this study and the rest of the team of the PSMAR screening technical office: Isabel Amatriain, Gloria Lagarriga, Maria Ángeles Mercader, Marina Reyes, Judit Silvilla and Eva Fernández.

## Supplementary file 1

### Crude logistic-regression models

**Model 1.** Crude participation.

| PARTICIPATION (POST-COVID-19) |  |  |  |  |  |  |  |  |
| --- | --- | --- | --- | --- | --- | --- | --- | --- |
|  | Estimate | Std. Error | z value | Pr(>\|z\|) | odds | OR post/pre | IC- (95%) | IC+ (95%) |
| FIRST-TIME INVITEE | -0.14765 | 0.03226 | -4.57 | 4.71e-06 *** | 0.862733019 | 0.862733019 | 0.8098203 | 0.9189847 |
| PREVIOUS NONPARTICIPANT | -2.06799 | 0.04017 | -51.4 | < 2e-16 *** | 0.12643967 | 1.08643443 | 1.00079898 | 1.178145 |
| REGULAR PARTICIPANT | 1.9568 | 0.02917 | 67.07 | < 2e-16 *** | 7.076645528 | 0.66289973 | 0.62686327 | 0.70126568 |
| IRREGULAR PARTICIPANT | -0.67188 | 0.04779 | -14.0 | < 2e-16 *** | 0.510747469 | 0.9574072 | 0.8671546 | 1.056191 |

**Model 2.** Crude Recall.

7 Model 2. Crude Recall.
| RECALL (POST-COVID-19) |  |  |  |  |  |  |  |  |
| --- | --- | --- | --- | --- | --- | --- | --- | --- |
|  | Estimate | Std. Error | z value | Pr(>\|z\|) | ODDS | OR post/pre | IC-(95%) | IC+(95%) |
| PREVALENT SCREENING | -0.10405 | 0.06385 | -1.63 | 0.103 | 0.901180237 | 0.901180237 | 0.7942011 | 1.0201312 |
| INCIDENT SCREENING | -1.51211 | 0.05957 | -25.3 | <2e-16 *** | 0.220444349 | 0.894956648 | 0.79856602 | 1.00038578 |

**Model 3.** Crude false positives.

9 Model 3. Crude false positives.
| FALSE POSITIVES (POST-COVID-19) |  |  |  |  |  |  |  |  |
| --- | --- | --- | --- | --- | --- | --- | --- | --- |
|  | Estimate | Std. Error | z value | Pr(>\|z\|) | ODDS | OR post/pre | IC-(95%) | IC+(95%) |
| PREVALENT SCREENING | 0.0883 | 0.07233 | 1.22 | 0.222 | 1.092337 | 1.09233761 | 0.94648207 | 1.25688409 |
| INCIDENT SCREENING | -1.4248 | 0.07139 | -19.95 | <2e-16 *** | 0.240534 | 0.94423519 | 0.82303271 | 1.0792094 |

**Model 4.** Crude cancer detection.

1 Model 4. Crude cancer detection.
| CANCER DETECTION (POST-COVID-19) |  |  |  |  |  |  |  |  |
| --- | --- | --- | --- | --- | --- | --- | --- | --- |
|  | **Estimate** | **Std. Error** | **z value** | **Pr(>\|z\|)** | **ODDS** | **OR post/pre** | **IC-(95%)** | **IC+(95%)** |
| PREVALENT SCREENING | -0.08015 | 0.28029 | -0.286 | 0.775 | 0.922977 | 0.922977 | 0.515085 | 1.55732 |
| INCIDENT SCREENING | -0.24311 | 0.18435 | -1.319 | 0.187 | 0.784185 | 0.893820 | 0.661616 | 1.18544 |

